# Impact of HLA divergence on humoral response to SARS-CoV-2 and HBV vaccines in the liver transplantation setting

**DOI:** 10.1101/2022.08.29.22279247

**Authors:** Cyrille Féray, Vincent Allain, Jean Luc Taupin, Bruno Roche, Christophe Desterke, Ilias Kounis, Zeynep Demir, Anne-Marie Roque-Afonso, Audrey Coilly, Didier Samuel, Sophie Caillat-Zucman

**Author notes:** equal contribution (Didier Samuel, Sophie Caillat-Zucman). Role of the funding source No funding external to the INSERM was provided for this study. The funding source had no role in the design or conduct of the study, analysis of the data, or the decision to submit the manuscript for publication.

## Abstract

**Background:** Organ transplant recipients are at high risk of viral infections but show lower humoral vaccine responsiveness than immunocompetent individuals. HLA evolutionary divergence (HED) quantifies the sequence differences between homologous HLA alleles and reflects the breadth of the immunopeptidome presented to T lymphocytes.

**Methods:** We retrospectively investigated the impact of HED on humoral response to SARS-CoV-2 mRNA vaccine in 310 liver transplant recipients (undetectable anti-spike IgG titers considered as no response, ≤250 BAU/mL as moderate response, >250 BAU/mL as strong response) and to Hepatitis B virus (HBV) vaccine in 424 liver transplant candidates (anti-HBs IgG <10 mIU/mL considered as no response, 10-100 mIU/mL as moderate reponse, ≥100 mIU/mL as strong response). HED between aligned allele pairs at HLA-A, -B, -DRB1 and- DQB1 loci were measured as a continuous metric using the Grantham distance. The impact of HED on vaccine responses was analyzed through ordinal logistic regression and inverse probability weighting approach based on generalised propensity scores.

**Findings:** For both vaccines, HED at the DQB1 locus, but not at other loci, was significantly higher in responders than in others, independent of covariates associated to the response (age, time since transplant, hemoglobin levels, combined graft, immunosuppression with steroids or mycophenolate for SARS-CoV-2 vaccine; age, gender, and liver disease for HBV vaccine).

**Interpretation:** DQB1 HED is a critical determinant of humoral response to vaccines in liver transplant recipients. This metric could guide the design of future vaccines as it predicts the magnitude of the repertoire of vaccine-derived peptides presented to CD4 helper T cells.

**Funding:** Institut National de la Sante et de la Recherche Medicale (INSERM)

## Introduction

Solid organ transplant recipients are at high risk of viral infections due to therapeutic immunosuppression. Therefore, pre-transplantation vaccination is strongly encouraged for vaccine-preventable diseases such as hepatitis B. Due to significantly increased morbidity and mortality from SARS-CoV-2 infection compared to the general population, organ transplant recipients were also rapidly prioritized in SARS-CoV-2 mRNA vaccination programs. However, these patients develop significantly impaired humoral responses compared to immunocompetent individuals ^1 2 3 4 5^.

Host-intrinsic factors such as age, gender, comorbidities, and genetics, particularly HLA polymorphisms, have been associated with vaccine responsiveness ^6^. HLA class 1 and class 2 molecules bind and present antigenic peptides (including vaccine-derived peptides) to CD8 and CD4 T lymphocytes, respectively, subsequently initiating the specific immune response. Thus, variations in the efficacy of the vaccine response may be related to the repertoire of HLA-bound peptides, called immunopeptidome.

At each HLA locus, the polymorphism is mostly concentrated in the exons encoding the peptide-binding groove of the corresponding HLA molecule. The divergent allele advantage hypothesis predicts that the more diverse the peptide-binding grooves of the 2 alleles of a given HLA gene, the wider the immunopeptidome presented by the corresponding HLA molecules, thereby increasing the probability of eliciting a broader immune response ^7^. Notably, HED emerged as a strong determinant of clinical outcomes in cancer patients treated with immune checkpoint inhibitors ^8 9 10 11 12 13 14 15^, although discrepant findings were reported in independent patient cohort ^16^. HED was also associated with overall survival in hematopoietic stem cell transplantation recipients and in patients with idiopathic bone marrow failure ^17 18 19^.

Studying a large cohort of liver transplant recipients with long-term follow-up and systematic liver biopsies, we recently demonstrated that a high class I HED of the donor is a strong and independent driver of allograft rejection, while HED of the recipient has no impact on the transplant outcome ^20^. Overall, these results demonstrate the relevance of HED as a predictor of the efficacy/strength of T cell responses in various settings. We hypothesized that HED could similarly impact the immunological response to vaccines. Taking advantage of systematic SARS-Cov2 and HBV vaccination programs in a cohort of liver transplant patients, we investigated the potential link between HED and the quality of humoral responses to these two vaccines.

## Patients and methods

The liver transplant recipient cohort, consisting of 909 adults and 113 children who received a liver transplant between January 2004 and January 2018 at Paul Brousse hospital or between 2010 and 2018 at Necker-Enfants Malades hospital, respectively, was recently described ^20^. HLA typing data were available in all patients. Immunosuppression consisted mainly of tacrolimus, tapered corticosteroids, and mycophenolate. In the event of kidney- or heart-combined transplantation, the immunosuppressive regimen used anti-thymoglobulins, higher doses of immunosuppressive drugs and a maintenance with corticosteroids.

The study was performed in accordance with the Declaration of Helsinki. Authorizations were given by the Local ethic committee (CPP bicetre N° CO 16-006) and the national commission for information technology and freedom (CNIL N° 1856085).

### SARS-Cov2 vaccine cohort

Between January and September 2021, 655 of the 909 liver transplant recipients (adults only) were included in the BNT162b2 mRNA vaccine program. At that time, a SARS-CoV-2 quantitative serological assay targeting the spike protein was planned 4 weeks after the second dose. A third dose was preferentially given to those who did not respond to the two injections, followed by a new serological assay 4 weeks later. Of the 655 vaccinated patients, 310 who had serology available 4 to 6 weeks after at least two injections were considered in this study, including 124 who received 2 doses and 186 who received 3 doses. Of the 186 patients who received 3 doses, 104 (56%) had no response after the second dose and therefore had a third injection. Patients with documented pre-vaccinal SARS-Cov2 infection or positive serology, or anticancer chemotherapy, were excluded. Flow chart is shown in Supplemental Figure 1A.

Response to SARS-CoV-2 vaccine was defined according to quantitative serological assays. The introduction of the WHO first International Standard for anti-SARS-CoV-2 immunoglobulin (NIBSC code 20/136) in December 2020 has allowed defining conversion factors (cf) from the arbitrary units (AU) of each manufacturer to Binding Antibody Units (BAU= cf x AU) to assist in comparing assays with the same protein specificity. Serological tests used in this study were the Elecsys Anti-SARS-CoV-2 S assay (Roche Diagnostics) and the SARS-CoV-2 IgG II Quant assay (Abbott Diagnostics), both targeting the RBD domain of the S1 subunit, and the LIAISON SARS-CoV-2 Trimeric S IgG assay (Diasorin) targeting the full Spike protein, with results reported in BAU/mL.

We adapted the correlates of protection of Feng et al., indicating that 80% protection against symptomatic infection was achieved with 264 (95% CI: 108, 806) BAU/mL measured 4 weeks after full vaccination ^21^. Therefore, an undetectable anti-S titer 4 weeks post-vaccination was considered as no response, detectable anti-S ≤250 BAU/ml as moderate response and > 250 BAU/mL as strong response. The 250 BAU threshold was used as it was the upper limit of linearity of the widely used Roche anti-S assay.

### HBV vaccine cohort

Before transplantation, 546 HBsAg- and HBc-negative patients received anti-HBV vaccination by ENGERIX (GSK) or GENHEVAC (Sanofi) during their transplant waiting time, with an accelerated vaccinal schedule (3 doses 1 month apart). Among them, 424 patients (including 78 children) who were fully vaccinated and had anti-HBs titration at 3 to 4 months after the third dose, were included in the present study. Flow Chart is given in Supplemental Figure 1B. HBV vaccine response was defined by anti-HBs titers as follows: <10 mIU/mL, no response, ≥ 10 but <100 mIU/mL, moderate response, and ≥100 mIU/mL, strong response.

### HLA data

HLA-A, -B, -DRB1 and -DQB1 typing at second field (four-digit) resolution was performed in all recipients and donors as reported ^20^. The respective protein sequences of the peptide-binding groove (exons 2 and 3 for HLA class I, and exon 2 for HLA class II) were extracted from the IMGT/HLA database ^22^. For each of the 4 HLA genes, the sequence divergence between 2 alleles (HED) was computed for all possible combinations of allele pairs encountered in the cohort. The calculation of HED between aligned allele pairs of a given locus was measured as a continuous metric using the Grantham distance ^7^, which takes into account the differences in the composition, polarity and volume of between two amino acids. By definition, HED is null in case of homozygosity. Importantly, HED data was not available in medical records and therefore did not influence any decisions regarding vaccination schedules.

### Statistical Analysis

Standard descriptive statistics were used to summarize baseline demographic, clinical and biological characteristics. For both vaccinations, the outcome was the vaccinal response defined as an ordinal categorial variable: no response, moderate response and strong response. Ordered logistic regression was used for the univariable and multivariable analysis of covariates (MASS package). The threshold of significance was p-value < 0.05).

HEDs are non-gaussian continuous metrics ^20^. To estimate the causal effect of HED as exposure on the vaccine response, we used the generalized propensity score (GPS) estimated via generalized boosted model (GBM). GBM estimates the inverse propensity score for the exposure using a flexible estimation method taking into account all covariates. GBM estimation involves an iterative process with multiple regression trees to capture relationships between the exposure and the covariates without over-fitting the data ^23^. For continuous exposures, average absolute Pearson correlation (AAC) between the exposure and each confounder was used to summarize balance performance at each iteration of the GBM fit, with the optimal iteration chosen as the one that minimizes the average or maximum AAC. An AAC with a cut-off value of 0.1 means that confounding effect of covariates is small ^23^. Finally, analysis of vaccine response as an ordinal outcome variable and exposure variable (HED) was done with a generalized linear model (GLM) using weights estimated by the GBM (Survey package).

For handling missing values, we applied the missing indicator method consisting of i) adding a missing data category for categorical covariates, and ii) setting the missing data to zero and adding supplementary binary covariates in the propensity score indicating for each continuous covariates whether the value is missing or not. When we had less than ten missing values on a covariate, we excluded the individuals to avoid convergence issues.

Associations of HLA phenotypic or allelic frequencies with the vaccine response were evaluated using a two-sided Fisher’s exact test with the Bonferroni correction.

All analyses were performed with R 4.0 ^24^ using the “ MASS”, “survey”, “publish”, “ggplot2”, “twangContinuous”, “survey”,”cobalt” and “Ggally” packages.

## Results

### SARS-CoV-2 vaccine response

Table 1 shows the characteristics of the 310 liver transplant recipients who received 2 or 3 doses of the BNT162b2 mRNA vaccine, according to the spike-specific antibody response defined by quantitative serological assays. Importantly, at the time of this study, the third dose was not yet systematic and was preferentially given to patients who had not responded to two doses. Altogether, 148 patients (47.5%) developed a strong response, 86 (27.5%) a moderate response, and 76 (25%) had no response. No serious adverse event was observed.

**Table 1.**
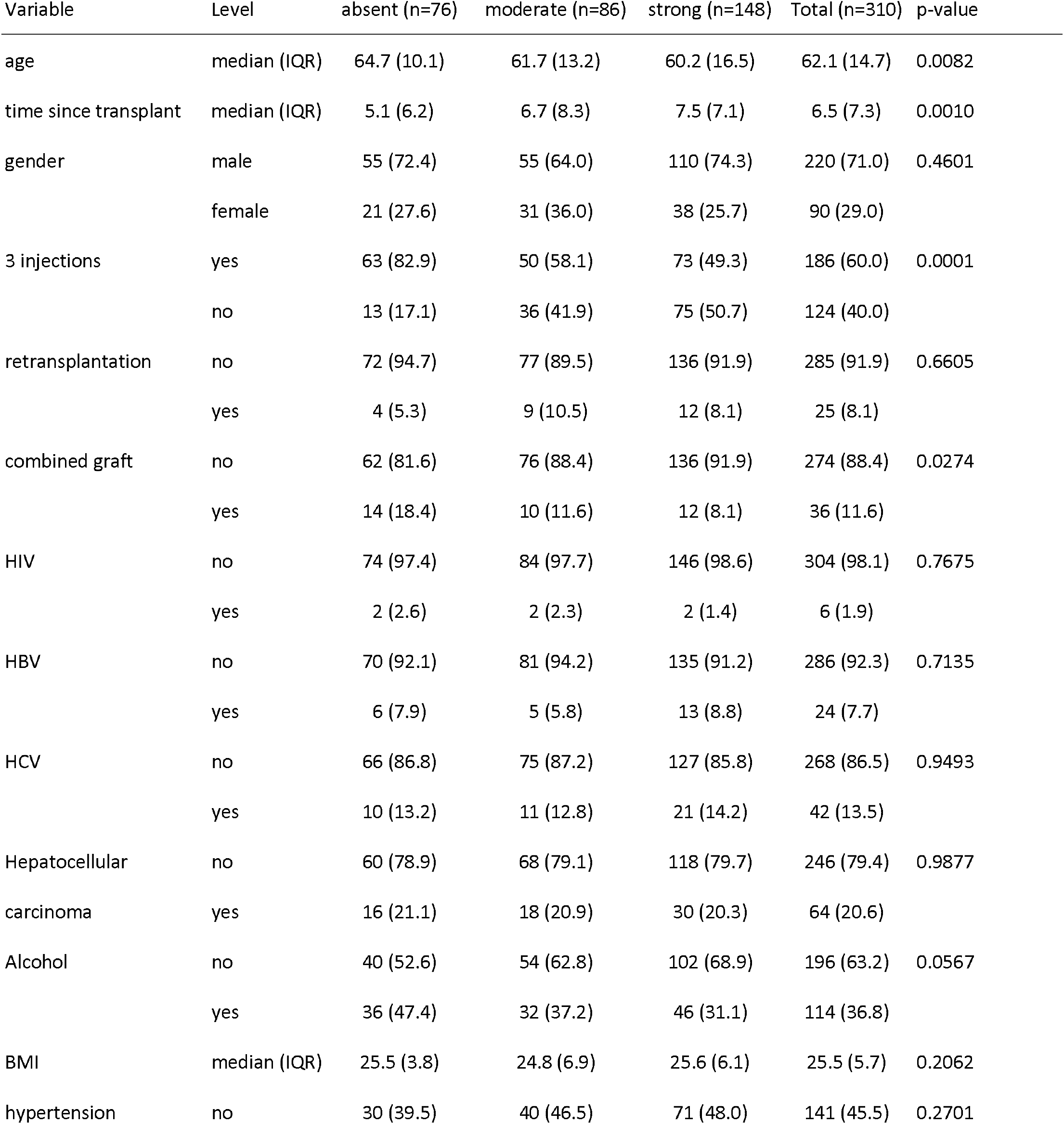
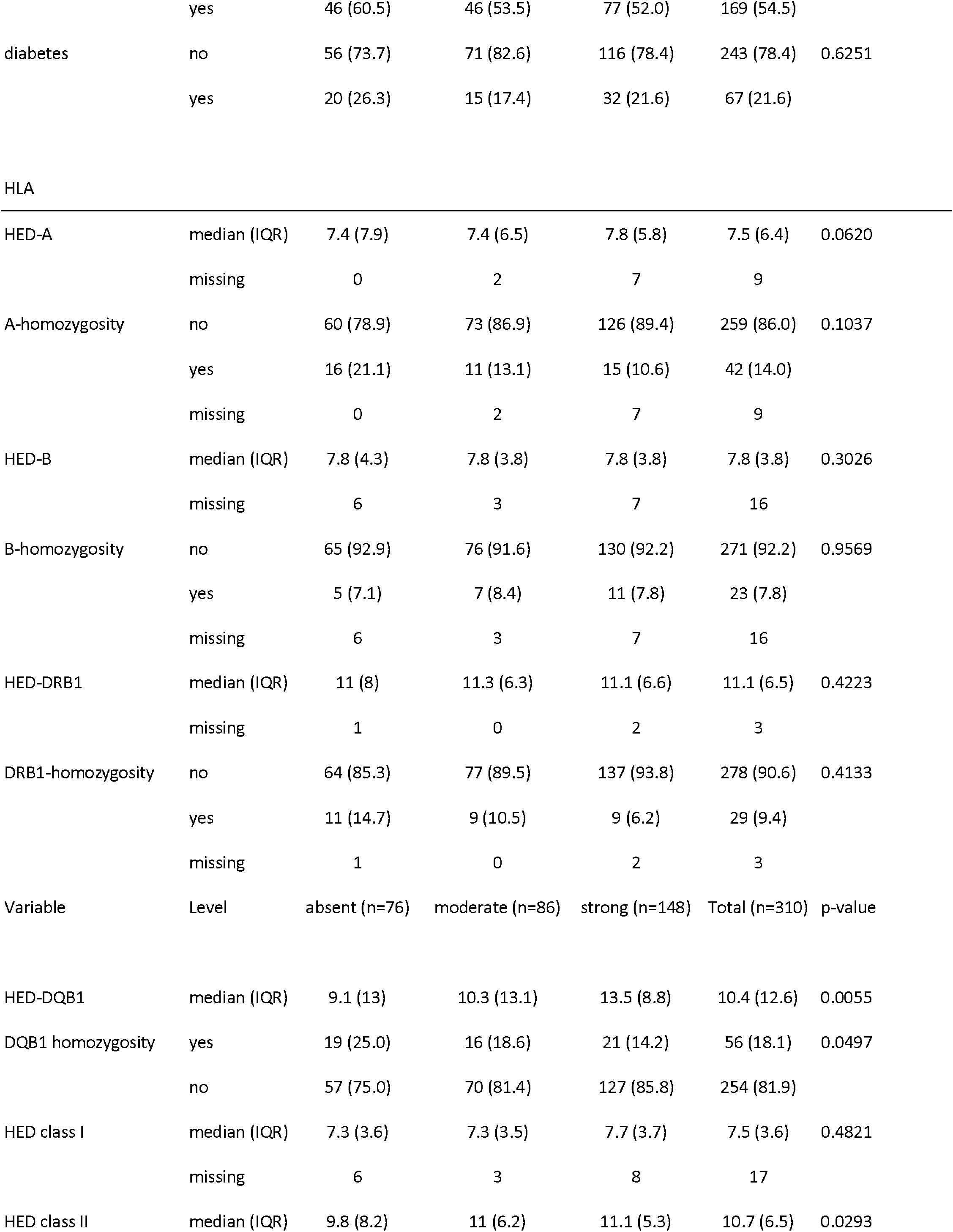
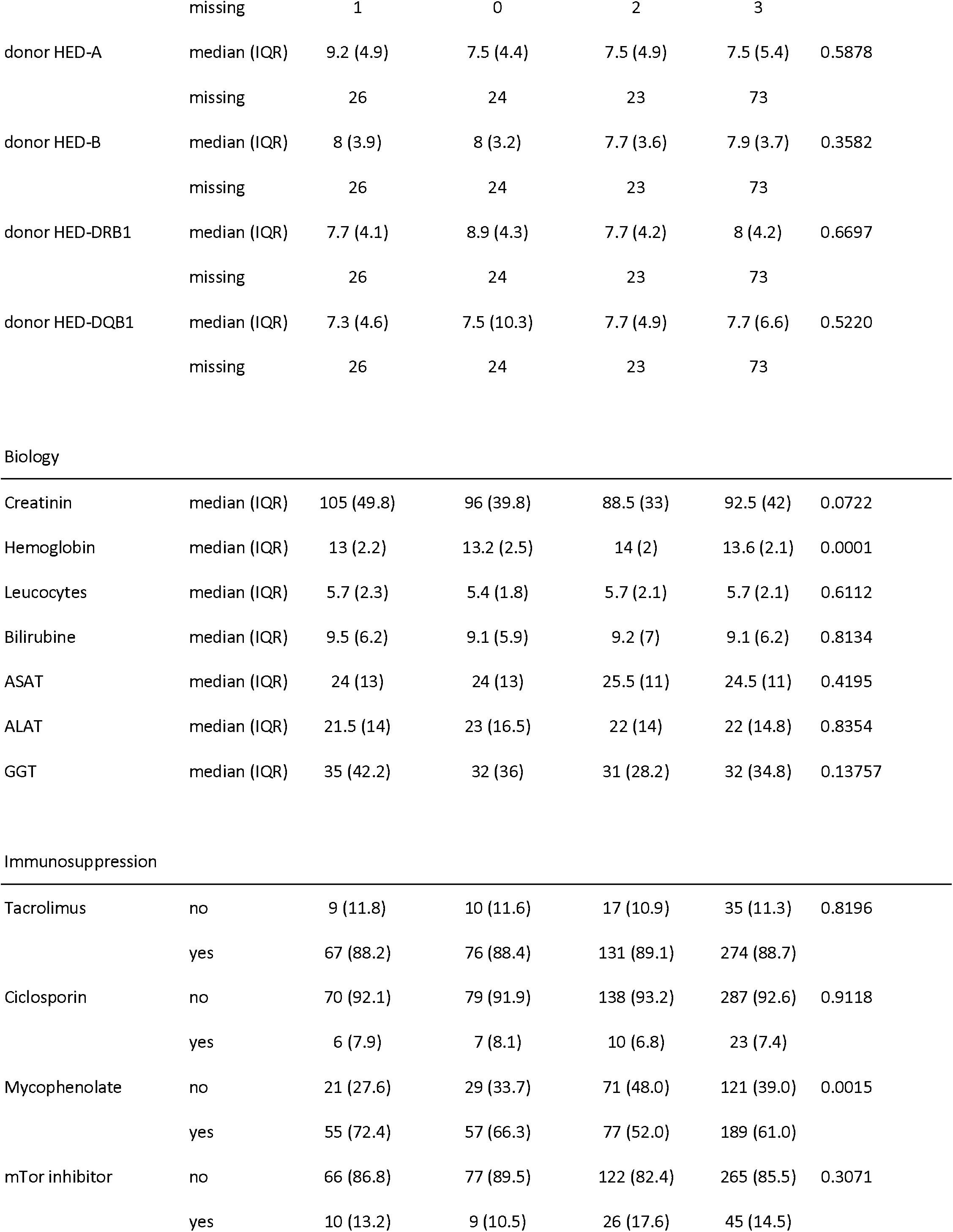
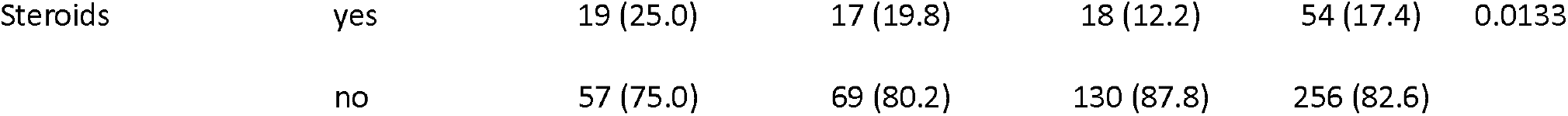
Univariate ordinal analysis of humoral response to SARS-CoV-2 vaccine

| Variable | Level | absent (n=76) | moderate (n=86) | strong (n=148) | Total (n=310) | p-value |
| --- | --- | --- | --- | --- | --- | --- |
| age | median (IQR) | 64.7 (10.1) | 61.7 (13.2) | 60.2 (16.5) | 62.1 (14.7) | 0.0082 |
| time since transplant | median (IQR) | 5.1 (6.2) | 6.7 (8.3) | 7.5 (7.1) | 6.5 (7.3) | 0.0010 |
| gender | male | 55 (72.4) | 55 (64.0) | 110 (74.3) | 220 (71.0) | 0.4601 |
|  | female | 21 (27.6) | 31 (36.0) | 38 (25.7) | 90 (29.0) |  |
| 3 injections | yes | 63 (82.9) | 50 (58.1) | 73 (49.3) | 186 (60.0) | 0.0001 |
|  | no | 13 (17.1) | 36 (41.9) | 75 (50.7) | 124 (40.0) |  |
| retransplantation | no | 72 (94.7) | 77 (89.5) | 136 (91.9) | 285 (91.9) | 0.6605 |
|  | yes | 4 (5.3) | 9 (10.5) | 12 (8.1) | 25 (8.1) |  |
| combined graft | no | 62 (81.6) | 76 (88.4) | 136 (91.9) | 274 (88.4) | 0.0274 |
|  | yes | 14 (18.4) | 10 (11.6) | 12 (8.1) | 36 (11.6) |  |
| HIV | no | 74 (97.4) | 84 (97.7) | 146 (98.6) | 304 (98.1) | 0.7675 |
|  | yes | 2 (2.6) | 2 (2.3) | 2 (1.4) | 6 (1.9) |  |
| HBV | no | 70 (92.1) | 81 (94.2) | 135 (91.2) | 286 (92.3) | 0.7135 |
|  | yes | 6 (7.9) | 5 (5.8) | 13 (8.8) | 24 (7.7) |  |
| HCV | no | 66 (86.8) | 75 (87.2) | 127 (85.8) | 268 (86.5) | 0.9493 |
|  | yes | 10 (13.2) | 11 (12.8) | 21 (14.2) | 42 (13.5) |  |
| Hepatocellular carcinoma | no | 60 (78.9) | 68 (79.1) | 118 (79.7) | 246 (79.4) | 0.9877 |
|  | yes | 16 (21.1) | 18 (20.9) | 30 (20.3) | 64 (20.6) |  |
| Alcohol | no | 40 (52.6) | 54 (62.8) | 102 (68.9) | 196 (63.2) | 0.0567 |
|  | yes | 36 (47.4) | 32 (37.2) | 46 (31.1) | 114 (36.8) |  |
| BMI | median (IQR) | 25.5 (3.8) | 24.8 (6.9) | 25.6 (6.1) | 25.5 (5.7) | 0.2062 |
| hypertension | no | 30 (39.5) | 40 (46.5) | 71 (48.0) | 141 (45.5) | 0.2701 |
|  | yes | 46 (60.5) | 46 (53.5) | 77 (52.0) | 169 (54.5) |  |
| diabetes | no | 56 (73.7) | 71 (82.6) | 116 (78.4) | 243 (78.4) | 0.6251 |
|  | yes | 20 (26.3) | 15 (17.4) | 32 (21.6) | 67 (21.6) |  |
| HLA |  |  |  |  |  |  |
| HED-A | median (IQR) | 7.4 (7.9) | 7.4 (6.5) | 7.8 (5.8) | 7.5 (6.4) | 0.0620 |
|  | missing | 0 | 2 | 7 | 9 |  |
| A-homozygosity | no | 60 (78.9) | 73 (86.9) | 126 (89.4) | 259 (86.0) | 0.1037 |
|  | yes | 16 (21.1) | 11 (13.1) | 15 (10.6) | 42 (14.0) |  |
|  | missing | 0 | 2 | 7 | 9 |  |
| HED-B | median (IQR) | 7.8 (4.3) | 7.8 (3.8) | 7.8 (3.8) | 7.8 (3.8) | 0.3026 |
|  | missing | 6 | 3 | 7 | 16 |  |
| B-homozygosity | no | 65 (92.9) | 76 (91.6) | 130 (92.2) | 271 (92.2) | 0.9569 |
|  | yes | 5 (7.1) | 7 (8.4) | 11 (7.8) | 23 (7.8) |  |
|  | missing | 6 | 3 | 7 | 16 |  |
| HED-DRB1 | median (IQR) | 11 (8) | 11.3 (6.3) | 11.1 (6.6) | 11.1 (6.5) | 0.4223 |
|  | missing | 1 | 0 | 2 | 3 |  |
| DRB1-homozygosity | no | 64 (85.3) | 77 (89.5) | 137 (93.8) | 278 (90.6) | 0.4133 |
|  | yes | 11 (14.7) | 9 (10.5) | 9 (6.2) | 29 (9.4) |  |
|  | missing | 1 | 0 | 2 | 3 |  |
| Variable | Level | absent (n=76) | moderate (n=86) | strong (n=148) | Total (n=310) | p-value |
| HED-DQB1 | median (IQR) | 9.1 (13) | 10.3 (13.1) | 13.5 (8.8) | 10.4 (12.6) | 0.0055 |
| DQB1 homozygosity | yes | 19 (25.0) | 16 (18.6) | 21 (14.2) | 56 (18.1) | 0.0497 |
|  | no | 57 (75.0) | 70 (81.4) | 127 (85.8) | 254 (81.9) |  |
| HED class I | median (IQR) | 7.3 (3.6) | 7.3 (3.5) | 7.7 (3.7) | 7.5 (3.6) | 0.4821 |
|  | missing | 6 | 3 | 8 | 17 |  |
| HED class II | median (IQR) | 9.8 (8.2) | 11 (6.2) | 11.1 (5.3) | 10.7 (6.5) | 0.0293 |

|  | missing | 1 | 0 | 2 | 3 |  |
| --- | --- | --- | --- | --- | --- | --- |
| donor HED-A | median (IQR) | 9.2 (4.9) | 7.5 (4.4) | 7.5 (4.9) | 7.5 (5.4) | 0.5878 |
|  | missing | 26 | 24 | 23 | 73 |  |
| donor HED-B | median (IQR) | 8 (3.9) | 8 (3.2) | 7.7 (3.6) | 7.9 (3.7) | 0.3582 |
|  | missing | 26 | 24 | 23 | 73 |  |
| donor HED-DRB1 | median (IQR) | 7.7 (4.1) | 8.9 (4.3) | 7.7 (4.2) | 8 (4.2) | 0.6697 |
|  | missing | 26 | 24 | 23 | 73 |  |
| donor HED-DQB1 | median (IQR) | 7.3 (4.6) | 7.5 (10.3) | 7.7 (4.9) | 7.7 (6.6) | 0.5220 |
|  | missing | 26 | 24 | 23 | 73 |  |
| Biology |  |  |  |  |  |  |
| Creatinin | median (IQR) | 105 (49.8) | 96 (39.8) | 88.5 (33) | 92.5 (42) | 0.0722 |
| Hemoglobin | median (IQR) | 13 (2.2) | 13.2 (2.5) | 14 (2) | 13.6 (2.1) | 0.0001 |
| Leucocytes | median (IQR) | 5.7 (2.3) | 5.4 (1.8) | 5.7 (2.1) | 5.7 (2.1) | 0.6112 |
| Bilirubine | median (IQR) | 9.5 (6.2) | 9.1 (5.9) | 9.2 (7) | 9.1 (6.2) | 0.8134 |
| ASAT | median (IQR) | 24 (13) | 24 (13) | 25.5 (11) | 24.5 (11) | 0.4195 |
| ALAT | median (IQR) | 21.5 (14) | 23 (16.5) | 22 (14) | 22 (14.8) | 0.8354 |
| GGT | median (IQR) | 35 (42.2) | 32 (36) | 31 (28.2) | 32 (34.8) | 0.13757 |
| Immunosuppression |  |  |  |  |  |  |
| Tacrolimus | no | 9 (11.8) | 10 (11.6) | 17 (10.9) | 35 (11.3) | 0.8196 |
|  | yes | 67 (88.2) | 76 (88.4) | 131 (89.1) | 274 (88.7) |  |
| Ciclosporin | no | 70 (92.1) | 79 (91.9) | 138 (93.2) | 287 (92.6) | 0.9118 |
|  | yes | 6 (7.9) | 7 (8.1) | 10 (6.8) | 23 (7.4) |  |
| Mycophenolate | no | 21 (27.6) | 29 (33.7) | 71 (48.0) | 121 (39.0) | 0.0015 |
|  | yes | 55 (72.4) | 57 (66.3) | 77 (52.0) | 189 (61.0) |  |
| mTor inhibitor | no | 66 (86.8) | 77 (89.5) | 122 (82.4) | 265 (85.5) | 0.3071 |
|  | yes | 10 (13.2) | 9 (10.5) | 26 (17.6) | 45 (14.5) |  |
| Steroids | yes | 19 (25.0) | 17 (19.8) | 18 (12.2) | 54 (17.4) | 0.0133 |
|  | no | 57 (75.0) | 69 (80.2) | 130 (87.8) | 256 (82.6) |  |

In univariable analysis with response as an ordinal dependent variable, higher age, a short duration of transplantation, a combined kidney graft, past alcoholism, high creatinine levels, low hemoglobin levels and immunosuppression using mycophenolate or steroids were associated with a lower response. As expected, the proportion of non-responders was high in patients who received 3 injections (63/186 patients, 34%), most of them having received the third dose because they had no response after the second. We observed a significant association of the DQB1 HED with vaccine response (median value, 13.5 in strong responders, 10.3 in moderate responders and 9.1 in non-responders, p= 0.0055). As shown in Figure 1A, DQB1 HED was higher in responders than in non-responders but had no significant impact on the magnitude (moderate or strong) of the response. DQB1 homozygosity was more frequent in case of low response, but neither HED nor homozygosity at other HLA loci had any significant effect (Table 1). As expected, donor HEDs had no effect (data not shown).

**Figure 1.**
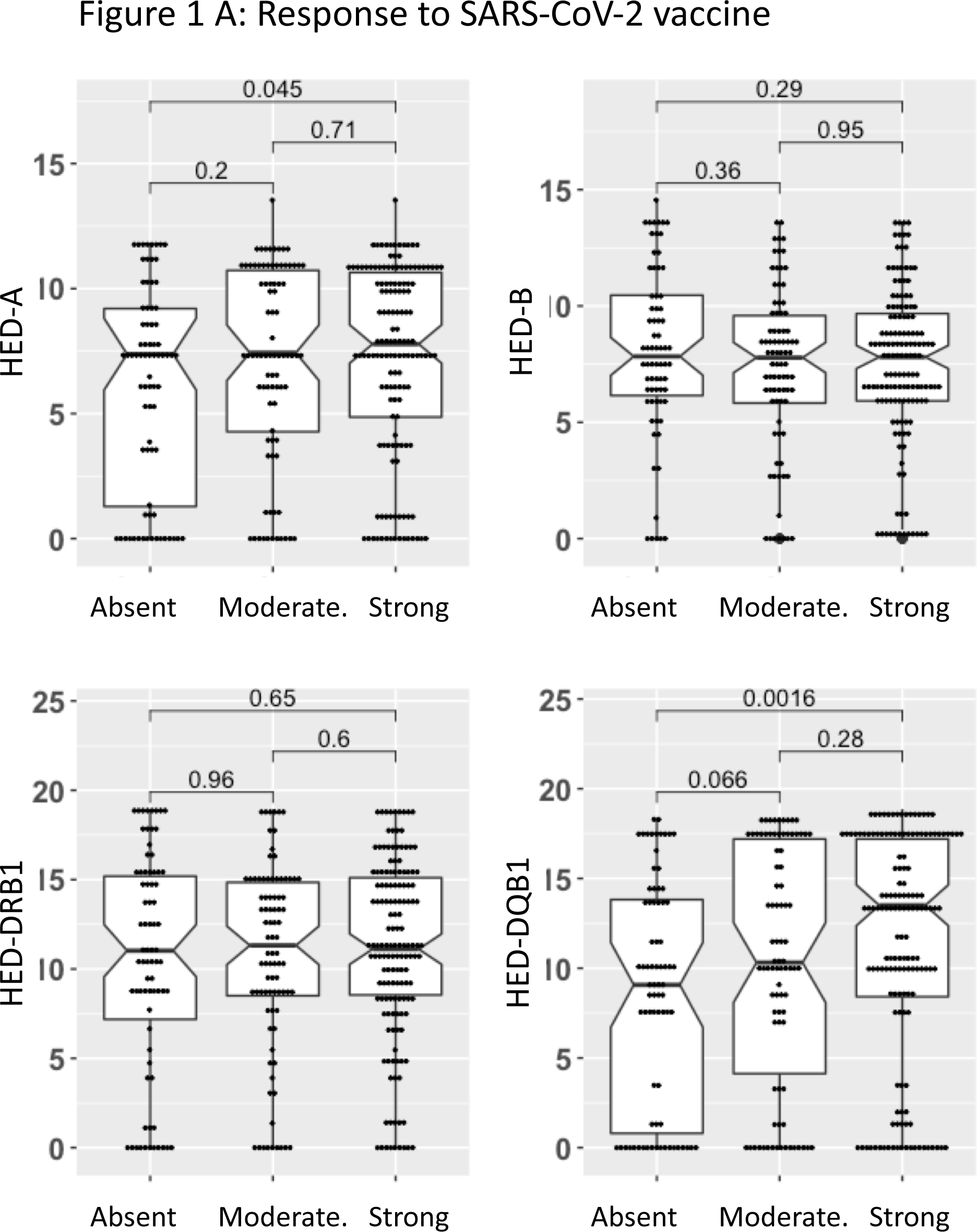
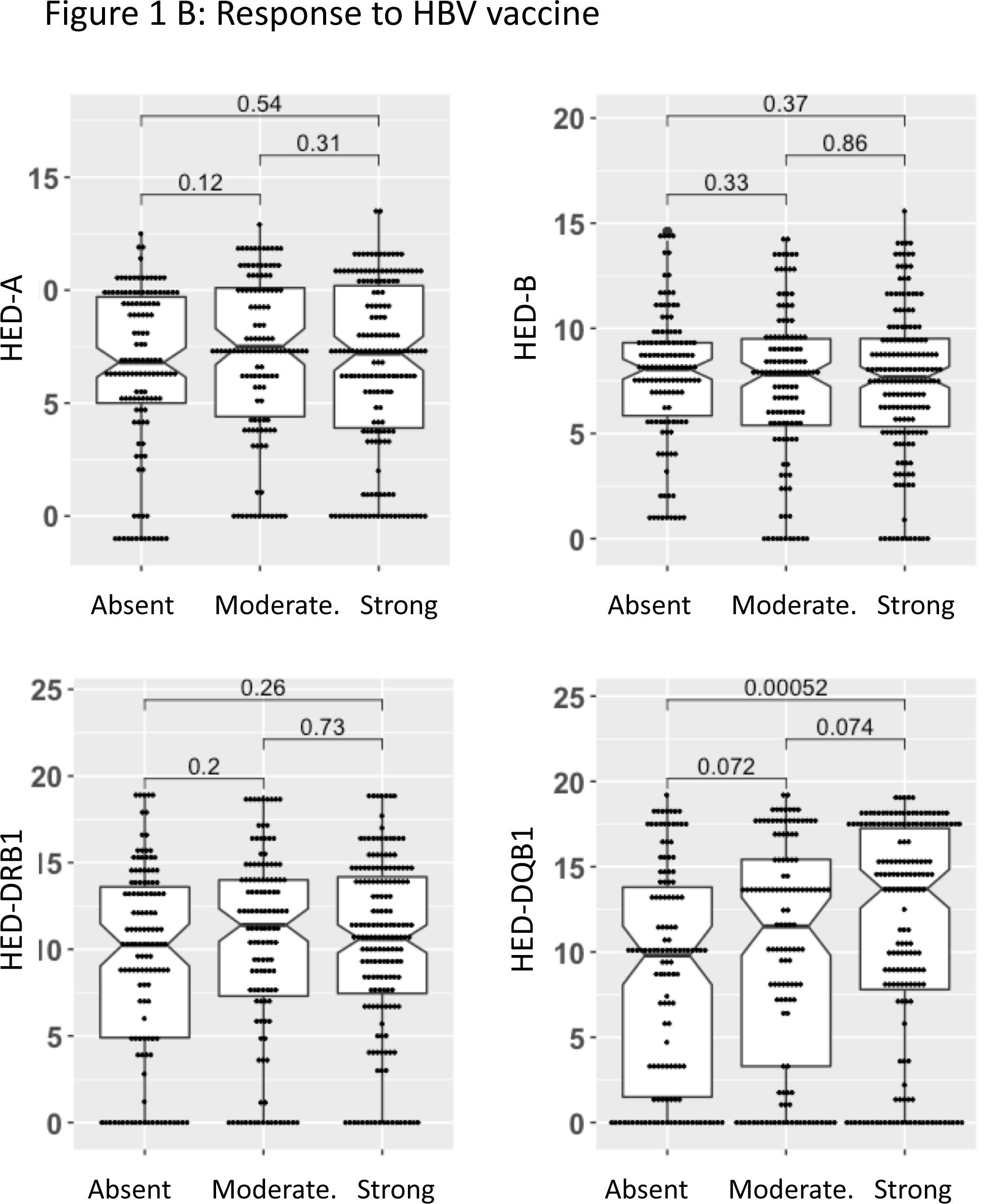
Boxplots of HED for HLA-A, -B, DRB1 and -DQB1 according to no, moderate or strong humoral response to SARS-CoV-2 (A, n= 310 patients) or HBV (B, n= 424 patients) vaccine. Two-sided Mann-Whitney P values are shown.

In multivariable ordinal logistic analysis including all the covariates which were significant in the univariate analysis except DQB1 homozygosity, the vaccine response was associated with DQB1 HED (odd ratio (OR)= 1.068, p= 0.0003), age, time since transplant, hemoglobin levels, steroids and mycophenolate (Table 2). A similar effect of the DQB1 HED was observed when analyzing separately the 186 patients who received three doses of vaccine (p= 0.0076) and the 124 patients who received only two doses (p= 0.0045) (Table 2). Similar results were observed when considering DQB1 homozygosity instead of DQB1 HED (Supplemental Table 1). However, the effect of DQB1 HED remained significant when the 50 DQB1 homozygous patients were excluded (Supplemental Table 2)

**Table 2.**
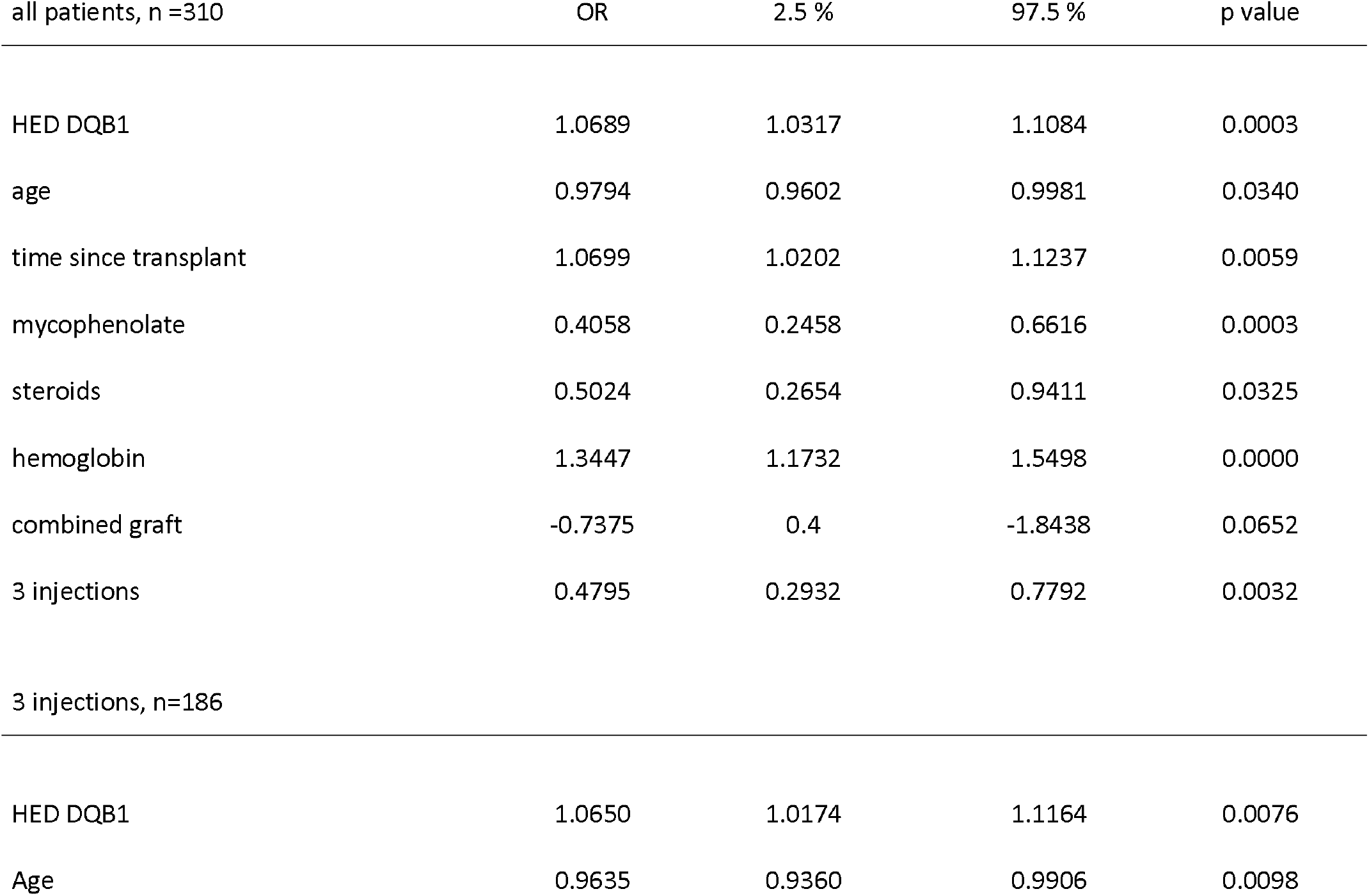
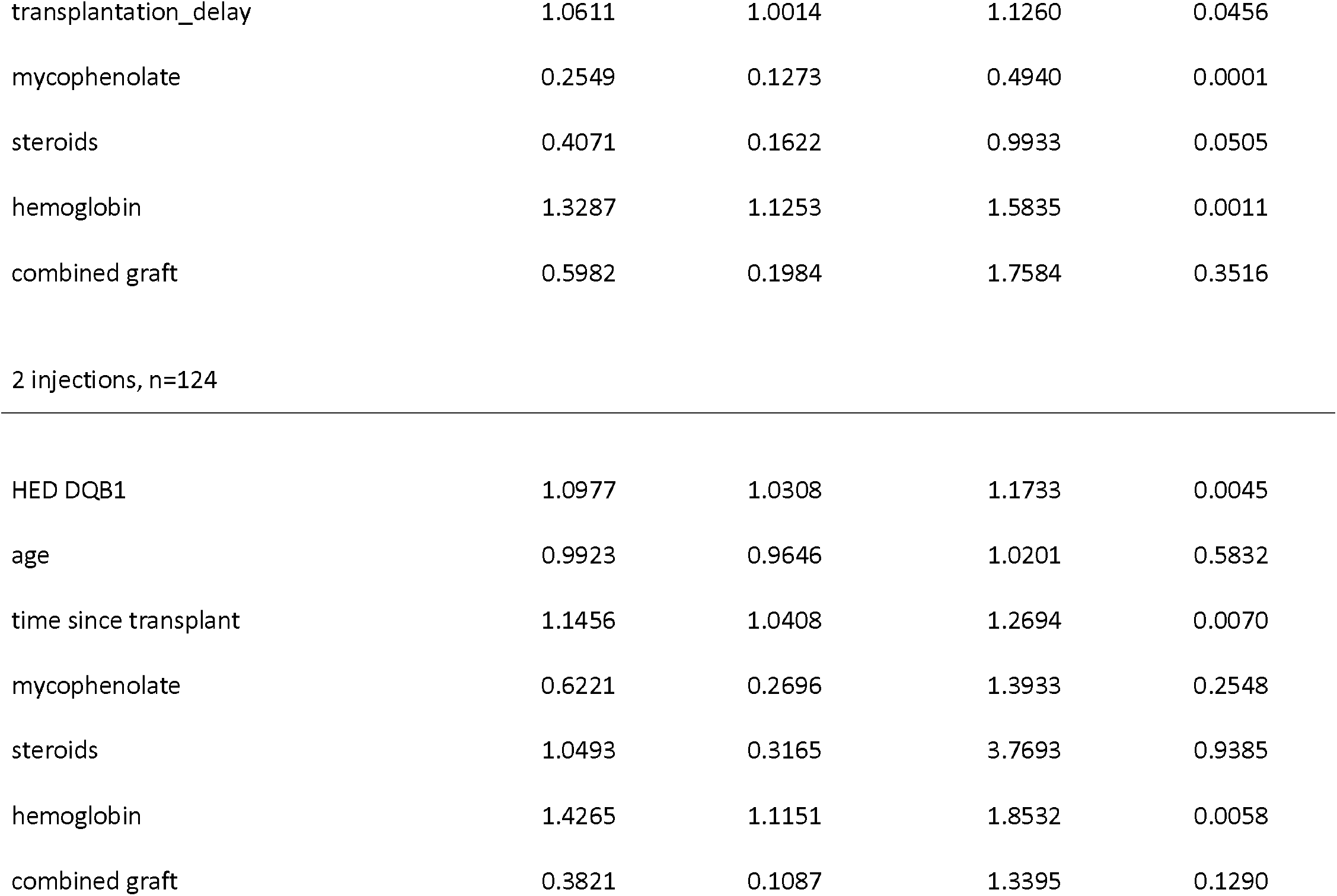
Multivariable Ordinal logistic regression of humoral response to SARS-CoV-2 vaccine

| all patients, n =310 | OR | 2.5 % | 97.5 % | p value |
| --- | --- | --- | --- | --- |
| HED DQB1 | 1.0689 | 1.0317 | 1.1084 | 0.0003 |
| age | 0.9794 | 0.9602 | 0.9981 | 0.0340 |
| time since transplant | 1.0699 | 1.0202 | 1.1237 | 0.0059 |
| mycophenolate | 0.4058 | 0.2458 | 0.6616 | 0.0003 |
| steroids | 0.5024 | 0.2654 | 0.9411 | 0.0325 |
| hemoglobin | 1.3447 | 1.1732 | 1.5498 | 0.0000 |
| combined graft | -0.7375 | 0.4 | -1.8438 | 0.0652 |
| 3 injections | 0.4795 | 0.2932 | 0.7792 | 0.0032 |
| 3 injections, n=186 |  |  |  |  |
| HED DQB1 | 1.0650 | 1.0174 | 1.1164 | 0.0076 |
| Age | 0.9635 | 0.9360 | 0.9906 | 0.0098 |
| transplantation_delay | 1.0611 | 1.0014 | 1.1260 | 0.0456 |
| mycophenolate | 0.2549 | 0.1273 | 0.4940 | 0.0001 |
| steroids | 0.4071 | 0.1622 | 0.9933 | 0.0505 |
| hemoglobin | 1.3287 | 1.1253 | 1.5835 | 0.0011 |
| combined graft | 0.5982 | 0.1984 | 1.7584 | 0.3516 |
| 2 injections, n=124 |  |  |  |  |
| HED DQB1 | 1.0977 | 1.0308 | 1.1733 | 0.0045 |
| age | 0.9923 | 0.9646 | 1.0201 | 0.5832 |
| time since transplant | 1.1456 | 1.0408 | 1.2694 | 0.0070 |
| mycophenolate | 0.6221 | 0.2696 | 1.3933 | 0.2548 |
| steroids | 1.0493 | 0.3165 | 3.7693 | 0.9385 |
| hemoglobin | 1.4265 | 1.1151 | 1.8532 | 0.0058 |
| combined graft | 0.3821 | 0.1087 | 1.3395 | 0.1290 |

Lastly, to precise the effect of HED as a continuous exposure, we used the generalized propensity score (GPS) estimated via the generalized boosted model (GBM), which is not sensitive to correlated covariates, thus allowing DQB1 homozygosity to be included as well. DQB1 HED was significantly associated with the vaccine response, independent of all other covariates (p = 0.021) (Supplemental Table 3).

### HBV vaccine cohort

The characteristics of the 424 patients who received a full anti-HBV vaccination before transplantation are shown in Table 3. As for the SARS-CoV-2 vaccine cohort, we analyzed the vaccine response according to anti-HBs titers 3 months after the end of the vaccination: 127 patients (30%) were non-responders, 126 (30%) had a moderate response, and 171 (40%) had a strong response. No serious adverse event was observed.

**Table 3.**
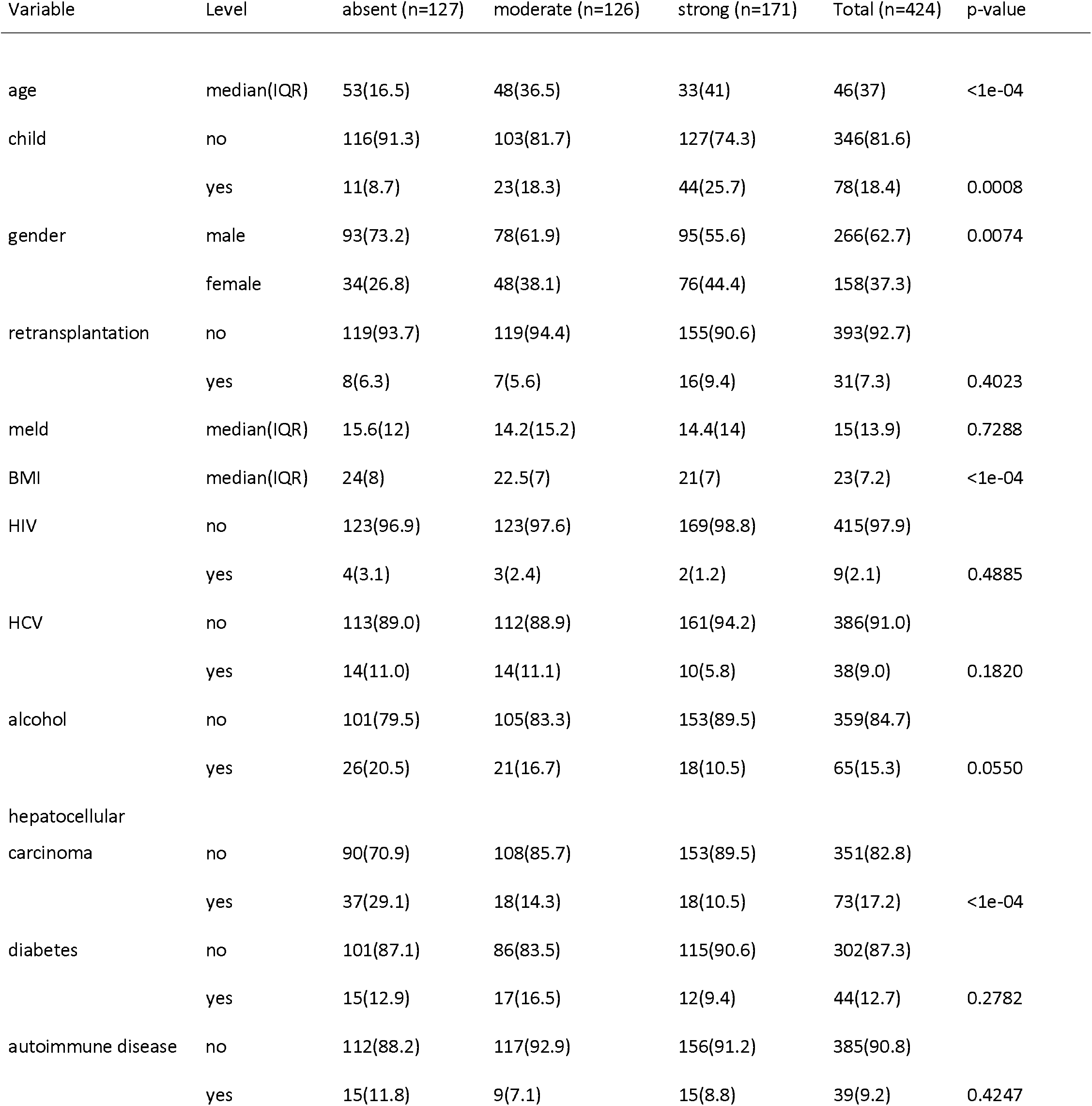
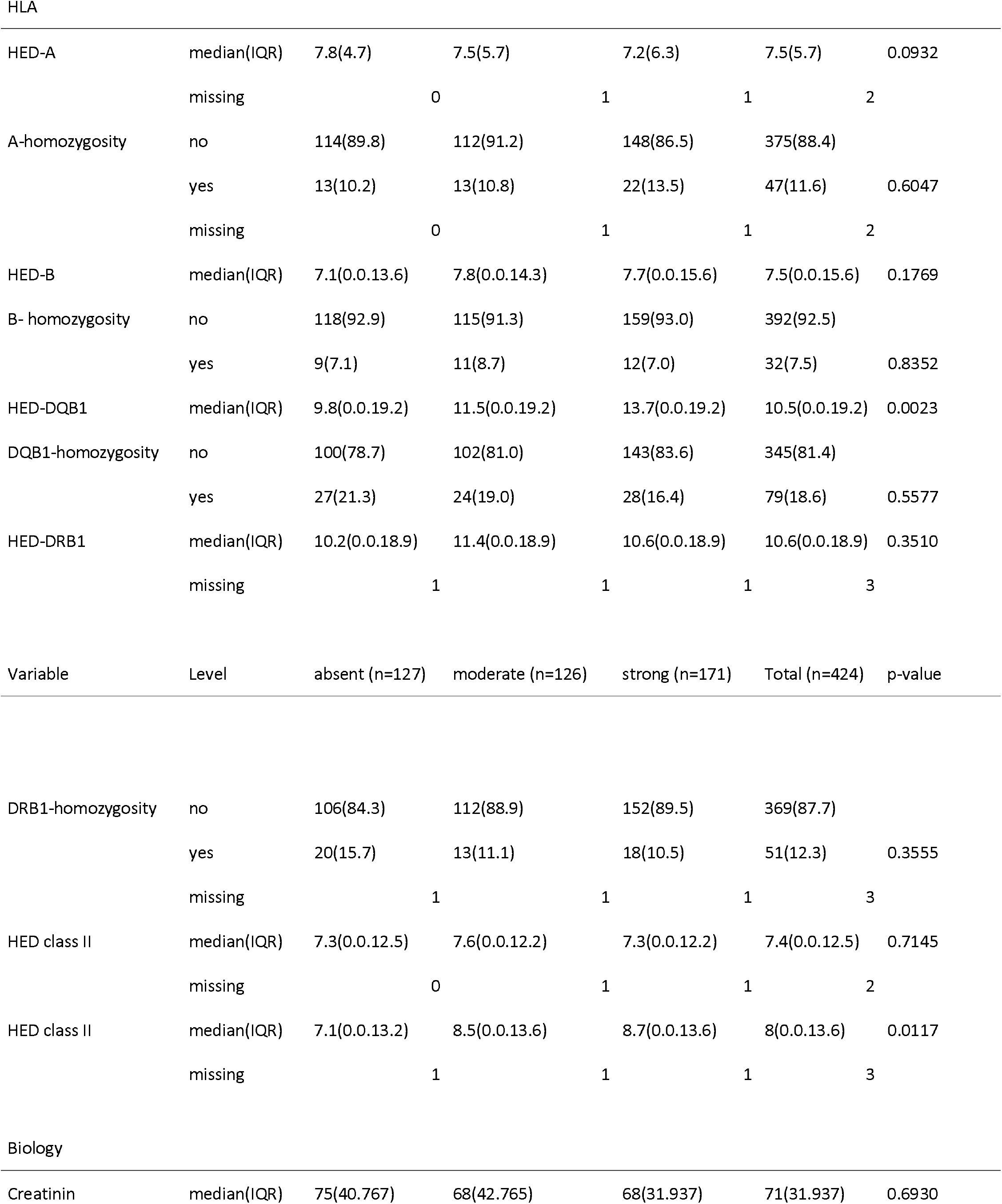
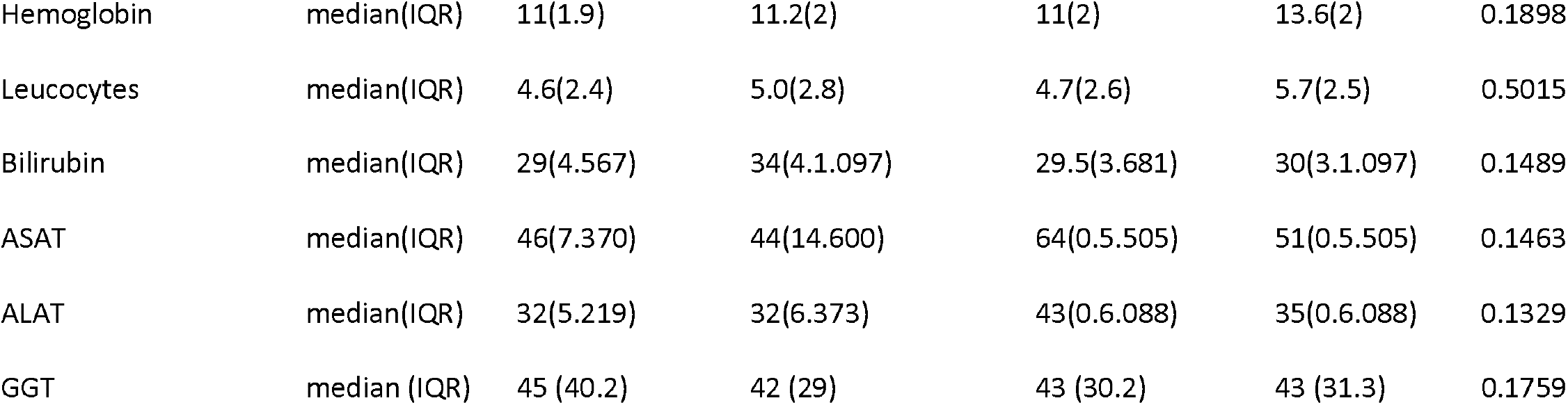
Univariate ordinal analysis of humoral response to HBV vaccine.

**Table 4.** Multivariable ordinal analysis of humoral response to HBV vaccine

|  | OR | 2.5 % | 97.5 % | p value |
| --- | --- | --- | --- | --- |
| HED-DQB1 | 1.05 | 1.02 | 1.08 | 0.0003 |
| Gender male | 0.66 | 0.45 | 0.96 | 0.0334 |
| Age | 0.97 | 0.96 | 0.98 | $< 10^{-5}$ |
| Hepatocellular carcinoma | 0.56 | 0.33 | 0.94 | 0.0312 |

Using logistic univariate analysis with response as an ordinal dependent variable, higher age, male gender, liver cancer at indication and higher BMI were associated with a lower HBV vaccine response. Similar to SARS-CoV-2 vaccine response, DQB1 HED was also significantly associated with HBV vaccine response (median value, 9.8 in non-responders, 11.5 in moderate responders, 13.7 in strong responders, p= 0.002). HED at other loci or homozygosity had no impact on response (Table 3). In multivariable ordinal logistic analysis taking the covariates which were significant in the univariate analysis, HBV vaccine response was associated with DQB1 HED (OR= 1.053, p= 0.0003), age, gender, and liver cancer as indication (Table 3). Using the generalized propensity score, we confirmed that DQB1 HED was associated with vaccine response, independent of other covariates (p= 0.019) (Supplemental Table 3).

### Vaccine response and HLA allele frequencies

In the SARS-CoV-2 cohort and HBV cohort, 71 and 74 different HLA alleles, respectively, were present in at least 5 patients. Phenotypic and allelic frequencies are shown in Supplemental Table 4. No HLA allele was associated with vaccine response to SARS-CoV-2 or HBV.

## Discussion

The response to vaccination is complex, likely involving the interplay of multiple and potentially confounding host factors. One mechanism associated with variations in vaccine response is the polymorphism of HLA genes. The exceptional HLA polymorphism, which was likely maintained by balancing selection under host-pathogen coevolution, mostly corresponds to a strong sequence divergence between peptide binding domains of alleles at a given HLA locus. The consequence is that more divergent allele pairs experience less overlap in the repertoire of bound antigenic peptides, and therefore can present a broader range of potential antigens for T cell recognition ^25^.

The clinical impact of HED has recently been highlighted in different immunopathological contexts such as anti-tumor response and allogeneic response. Here, we highlight for the first time the role of HED in a more physiological setting that is the response to vaccines. Taking advantage of a cohort of liver transplant recipients in which we recently demonstrated the impact of donor class 1 HED on allograft rejection, we analyzed humoral responses induced by two different types of vaccine, the recent BNT162b2 mRNA-based SARS-Cov2 vaccine, and the old recombinant HBV surface antigen-based vaccine. We show that patients who did not develop significant antibody response following 2 or 3 doses of the BNT162b2 mRNA vaccine carry HLA-DQB1 alleles with reduced sequence divergence between their peptide binding domains, not explained by a higher frequency of homozygosity. Importantly, this observation holds true for the HBV vaccine based on recombinant viral protein subunit. Although the mechanisms by which these different vaccine strategies induce a protective humoral response may be somewhat different, our results suggest that the more divergent the patient’s DQB1 molecules, the more diverse the vaccine-derived peptides presented to CD4 T helper cells, and the more efficient the B cell response and antibody production. Indeed, most protective antibody response are dependent on CD4 T-cell help, as demonstrated in the context of SARS-Cov2 infection by the correlation between spike-specific CD4 T cell response and the magnitude of anti-spike IgG titers^26^.

An intriguing question is the impact of divergence between peptide binding domains of DQB1 alleles, but not of DRB1 alleles. These two genes are located at adjacent loci in the HLA class II region and are in strong linkage disequilibrium, but DQB1 is much less polymorphic, i.e. exhibits a smaller number of different alleles, than DRB1. Whether this lower polymorphism can be compensated by a higher sequence divergence between DQB1 alleles is unclear. Moreover, HLA class II molecules consist of an αβ heterodimer encoded by the respective A and B genes at each locus, and both α and β chains contribute to the peptide binding domain of the molecule. For the DR molecules, the DRA gene is not polymorphic and therefore does not contribute to the overall divergence. This is probably different for DQ molecules because the DQA1 gene itself is moderately polymorphic and could participate to an additional layer of diversity in the peptide-binding groove. We were unable to verify this hypothesis because DQA1 genotyping is not routinely performed in the context of solid organ transplantation.

Independent of the effect of the DQB1 HED, our study confirms the role of factors previously associated with SARS-Cov2 vaccine response in liver transplant recipients, such as age, time since transplant, mycophenolate treatment^27^, and identifies others not described, such as the hemoglobin level. Data on HBV vaccine response in liver transplant candidates are scarce in the literature. The role of age, gender and chronic liver disease has been reported^6^, but not that of hepatocellular carcinoma which we observe.

This study has several limitations. First, surrogate markers of vaccine-induced protection are imperfect, and vaccine response is far more complex than measurable antibody titers. For SARS-Cov2 vaccine, we quantified anti-spike humoral response using commercial serologic tests rather than formal neutralization capacity, not available at this time. However, a strong correlation between anti-S IgG titer and neutralization capacity has been demonstrated, including in organ transplant recipients ^28 29 30 31^. Second, pre-vaccination SARS-Cov2 serology was only performed in one third of patients. However, the seroprevalence was very low (7/202 patients, 3%) at this time of the pandemic. Last, we did not directly test the hypothesis that reduced HED correlates with a less diverse repertoire of vaccine-derived peptides presented by HLA-DQ molecules. Thus, future studies to investigate causal relationships between these parameters are needed.

In conclusion, our study highlights HLA-DQ diversity as critical for vaccine responsiveness in liver transplant recipients. The consistency of the results with the two vaccines suggests that it is likely a broader mechanism also concerning immunocompetent individuals, although this remains to be confirmed. These results could guide the design of improved vaccine strategies most likely to elicit protective immunity, by predicting the magnitude of the repertoire of vaccine-derived peptides bound to HLA-DQ molecules.

## Data Availability

All data produced in the present study are available upon reasonable request to the authors

## Abbreviations

GBM: Gradient Boosted Models
HBV: Hepatitis B Virus
HCV: Hepatitis C Virus
HED: HLA Evolutionary Divergence
HIV: Human Immunodeficiency Virus
HLA: Human Leucocyte Antigen
IPW: Inverse Probability Weighting

## Acknowledgments

we are deeply grateful to Coralie Chopineaux, Alexandra Kokar, Gladys Saulac and Gaël Berthelot who organized the vaccination follow-up and collected the data.

## Notes

### Competing Interest Statement

The authors have declared no competing interest.

### Funding Statement

This study was funded by INSERM

## References

1. Thuluvath PJ, Robarts P, Chauhan M. Analysis of antibody responses after COVID-19 vaccination in liver transplant recipients and those with chronic liver diseases. J Hepatol 2021;75(6):1434–9.

2. Rincon-Arevalo H, Choi M, Stefanski A-L, et al. Impaired humoral immunity to SARS-CoV-2 BNT162b2 vaccine in kidney transplant recipients and dialysis patients. Sci Immunol 2021;6(60):eabj1031.

3. Sattler A, Schrezenmeier E, Weber UA, et al. Impaired humoral and cellular immunity after SARS-CoV-2 BNT162b2 (tozinameran) prime-boost vaccination in kidney transplant recipients. J Clin Invest 2021;131(14):150175.

4. Boyarsky BJ, Werbel WA, Avery RK, et al. Antibody Response to 2-Dose SARS-CoV-2 mRNA Vaccine Series in Solid Organ Transplant Recipients. JAMA 2021;325(21):2204.

5. Mitchell J, Connolly CM, Chiang TP-Y, et al. Comparison of SARS-CoV-2 Antibody Response After 2-Dose mRNA-1273 vs BNT162b2 Vaccines in Incrementally Immunosuppressed Patients. JAMA Netw Open 2022;5(5):e2211897.

6. Zimmermann P, Curtis N. Factors That Influence the Immune Response to Vaccination. Clin Microbiol Rev 2019;32(2):e00084–18.

7. Pierini F, Lenz TL. Divergent Allele Advantage at Human MHC Genes: Signatures of Past and Ongoing Selection. Mol Biol Evol 2018;35(9):2145–58.

8. Chowell D, Morris LGT, Grigg CM, et al. Patient HLA class I genotype influences cancer response to checkpoint blockade immunotherapy. Science 2018;359(6375):582–7.

9. Chowell D, Krishna C, Pierini F, et al. Evolutionary divergence of HLA class I genotype impacts efficacy of cancer immunotherapy. Nat Med 2019;25(11):1715–20.

10. Chowell D, Yoo S-K, Valero C, et al. Improved prediction of immune checkpoint blockade efficacy across multiple cancer types. Nat Biotechnol [Internet] 2021 [cited 2021 Nov 14];Available from: https://www.nature.com/articles/s41587-021-01070-8

11. Lee C-H, DiNatale RG, Chowell D, et al. High Response Rate and Durability Driven by HLA Genetic Diversity in Patients with Kidney Cancer Treated with Lenvatinib and Pembrolizumab. Mol Cancer Res 2021;19(9):1510–21.

12. Delyon J, Biard L, Renaud M, et al. PD-1 blockade with pembrolizumab in classic or endemic Kaposi’s sarcoma: a multicentre, single-arm, phase 2 study. The Lancet Oncology 2022;23(4):491–500.

13. Lu Z, Chen H, Jiao X, et al. Germline HLA-B evolutionary divergence influences the efficacy of immune checkpoint blockade therapy in gastrointestinal cancer. Genome Med 2021;13(1):175.

14. Manczinger M, Koncz B, Balogh GM, et al. Negative trade-off between neoantigen repertoire breadth and the specificity of HLA-I molecules shapes antitumor immunity. Nat Cancer 2021;2(9):950–61.

15. Litchfield K, Reading JL, Puttick C, et al. Meta-analysis of tumor- and T cell-intrinsic mechanisms of sensitization to checkpoint inhibition. Cell 2021;184(3):596–614.e14.

16. Chhibber A, Huang L, Zhang H, et al. Germline HLA landscape does not predict efficacy of pembrolizumab monotherapy across solid tumor types. Immunity 2022;55(1):56–64.e4.

17. Roerden M, Nelde A, Heitmann JS, et al. HLA Evolutionary Divergence as a Prognostic Marker for AML Patients Undergoing Allogeneic Stem Cell Transplantation. Cancers 2020;12(7):1835.

18. Daull A-M, Dubois V, Labussière-Wallet H, et al. Class I/Class II HLA Evolutionary Divergence Ratio Is an Independent Marker Associated With Disease-Free and Overall Survival After Allogeneic Hematopoietic Stem Cell Transplantation for Acute Myeloid Leukemia. Front Immunol 2022;13:841470.

19. Pagliuca S, Gurnari C, Awada H, et al. The similarity of class II HLA genotypes defines patterns of autoreactivity in idiopathic bone marrow failure disorders. Blood 2021;138(26):2781–98.

20. Féray C, Taupin J-L, Sebagh M, et al. Donor HLA Class 1 Evolutionary Divergence Is a Major Predictor of Liver Allograft Rejection[: A Retrospective Cohort Study. Ann Intern Med 2021;174(10):1385–94.

21. Feng S, Phillips DJ, White T, et al. Correlates of protection against symptomatic and asymptomatic SARS-CoV-2 infection. Nat Med 2021;27(11):2032–40.

22. Robinson J, Halliwell JA, Hayhurst JD, Flicek P, Parham P, Marsh SGE. The IPD and IMGT/HLA database: allele variant databases. Nucleic Acids Res 2015;43(Database issue):D423–431.

23. Zhu Y, Coffman DL, Ghosh D. A Boosting Algorithm for Estimating Generalized Propensity Scores with Continuous Treatments. J Causal Inference 2015;3(1):25–40.

24. ano ano. R Development Core Team. R: A Language and Environment for Statistical Computing [Internet]. Computing RF for S, editor. Vienna, Austria; 2010.http://www.R-project.org/.

25. Lenz TL. Computational prediction of MHC II-antigen binding supports divergent allele advantage and explains trans-species polymorphism. Evolution 2011;65(8):2380–90.

26. Grifoni A, Weiskopf D, Ramirez SI, et al. Targets of T Cell Responses to SARS-CoV-2 Coronavirus in Humans with COVID-19 Disease and Unexposed Individuals. Cell 2020;181(7):1489–1501.e15.

27. Toniutto P, Cussigh A, Cmet S, et al. Immunogenicity and safety of a third dose of anti-SARS-CoV-2 BNT16b2 vaccine in liver transplant recipients. Liver Int 2022;

28. Doffizi G, Agrati C, Visco-Comandini U, et al. Coordinated cellular and humoral immune responses after two-dose SARS-CoV2 mRNA vaccination in liver transplant recipients. Liver International 2022;42(1):180–6.

29. Charmetant X, Espi M, Benotmane I, et al. Infection or a third dose of mRNA vaccine elicits neutralizing antibody responses against SARS-CoV-2 in kidney transplant recipients. Sci Transl Med 2022;14(636):eabl6141.

30. Manenti A, Gianchecchi E, Dapporto F, et al. Evaluation and correlation between SARS-CoV-2 neutralizing and binding antibodies in convalescent and vaccinated subjects. Journal of Immunological Methods 2022;500:113197.

31. Gilbert PB, Montefiori DC, McDermott AB, et al. Immune correlates analysis of the mRNA-1273 COVID-19 vaccine efficacy clinical trial. Science 2022;375(6576):43–50.

