## Supplemental for "Impact of HLA divergence on humoral response to SARS-CoV-2 and HBV vaccines in the liver transplantation setting"

Supplemental Figure 1

A. SARS-CoV2 vaccination cohort

Death before 1/1/2021: 181

No vaccination: 38

No available anti-Spike titration: 109

SARS-Cov2 Infection before or during vaccination: 24

Inapropriate serology ( <3 or > 6 weeks after last injection): 56

Only one injection: 149

B. HBV vaccination cohort

No vaccination: 212

No available anti-HBs titration: 32

Anti-HBc positive: 122

Transplantation before complete vaccination: 90

909 liver transplant patients with HLA genotyping

1022 candidates for liver transplantation with HLA genotyping

lost: 35

655 patients receiving one dose of BNT162b2 vaccine

310 Included patients

546 candidates receiving one dose of HBV vaccine

Previous vaccination: 142

424 Included patients

SARS-Cov2 positive serology before vaccination: 6/202 tested patients

### Supplemental Table 1

Ordered logistic regression of humoral response to SARS-CoV2 vaccine with DQB1 homozygosity instead of DQB1 HED

|  | **OR** | **2.5 %** | **97.5 %** | **P-value** |
| --- | --- | --- | --- | --- |
| DQB1 homozygosity | 0.48 | 0.27 | 0.85 | 0.0119 |
| age | 0.98 | 0.96 | 1.00 | 0.0208 |
| time since transplant | 1.08 | 1.04 | 1.14 | 0.0006 |
| mycophenolate | 0.39 | 0.24 | 0.63 | 0.0001 |
| steroids | 0.63 | 0.33 | 1.21 | 0.1626 |
| Hemoglobin levels | 1.30 | 1.14 | 1.48 | 0.0001 |
| combined graft | 0.45 | 0.21 | 0.97 | 0.0400 |

### Supplemental Table 2

Ordered logistic regression of humoral response to SARS-CoV2 vaccine in HLA-DQB1 heterozygous patients

|  | **OR** | **2.5 %** | **97.5 %** | **P-value** |
| --- | --- | --- | --- | --- |
| HED-DQB1 | 1.08 | 1.02 | 1.13 | 0.0053 |
| age | 0.98 | 0.96 | 0.99 | 0.0091 |
| time since transplant | 1.07 | 1.02 | 1.13 | 0.0100 |
| mycophenolate | 0.35 | 0.20 | 0.60 | 0.0001 |
| steroids | 0.66 | 0.31 | 1.42 | 0.2883 |
| Hemoglobin levels | 1.29 | 1.11 | 1.51 | 0.0001 |
| combined graft | 0.51 | 0.21 | 1.26 | 0.1419 |

### Supplemental Table 3

DQB1 HED and humoral response to SARS-CoV2 or HBV vaccines. Characteristic of the general propensity score (GPS) based on generalized boosted model (GBM): number of iterations. effective sample size. corrections by GPS and statistical t- and p-values of the generalized linear model (GLM).

|  |  | GBM |  | GPS |  |  |  | GLM |  |
| --- | --- | --- | --- | --- | --- | --- | --- | --- | --- |
|  |  | Iterations | Effective sample size | unweighted | | weighted |  | t-value | p-value |
|  | Exposure |  |  | max.AAC | mean.ACC | max.AAC | mean.AAC | |  |
| SARS-CoV2 | **HED DQB1** | **435** | **280/310** | **0.350** | **0.063** | **0.097** | **0.060** | **2.261** | **0.021** |
| HBV | **HED DQB1** | **1209** | **256/424** | **0.375** | **0.081** | **0.087** | **0.042** | **2.342** | **0.019** |

### Supplemental Table 4.

Phenotypic and allelic frequencies of HLA genotypes according to humoral response (absent versus moderate or strong). Only HLA alleles found in more than 5 patients were considered.

| SARS-CoV-2 | Phenotypic frequency | |  | Allelic frequency |  |  |
| --- | --- | --- | --- | --- | --- | --- |
|  | Response |  |  | Response |  |  |
|  | **Positive** | **Negative** | **p-value** | **Positive** | **Negative** | **p-value** |
| **A* 01:01** | 0.76 | 0.75 | 1.00 | 0.76 | 0.75 | 1.00 |
| **A* 02:01** | 0.71 | 0.79 | 0.29 | 0.68 | 0.78 | 0.04 |
| **A* 03:01** | 0.73 | 0.76 | 0.92 | 0.72 | 0.76 | 0.87 |
| **A* 11:01** | 0.85 | 0.74 | 0.48 | 0.85 | 0.75 | 0.50 |
| **A* 23:01** | 0.71 | 0.76 | 1.00 | 0.71 | 0.76 | 1.00 |
| **A* 24:02** | 0.85 | 0.73 | 0.19 | 0.84 | 0.75 | 0.31 |
| **A* 25:01** | 0.93 | 0.75 | 0.47 | 0.93 | 0.75 | 0.48 |
| **A* 26:01** | 0.71 | 0.76 | 0.98 | 0.73 | 0.76 | 1.00 |
| **A* 29:02** | 0.83 | 0.75 | 0.85 | 0.83 | 0.75 | 0.80 |
| **A* 30:01** | 0.73 | 0.76 | 1.00 | 0.75 | 0.75 | 1.00 |
| **A* 30:02** | 0.50 | 0.76 | 0.31 | 0.50 | 0.76 | 0.32 |
| **A* 31:01** | 0.74 | 0.76 | 1.00 | 0.74 | 0.76 | 1.00 |
| **A* 32:01** | 0.81 | 0.75 | 0.97 | 0.83 | 0.75 | 0.88 |
| **A* 33:01** | 0.90 | 0.75 | 0.78 | 0.90 | 0.75 | 0.78 |
| **A* 68:01** | 0.68 | 0.76 | 0.90 | 0.70 | 0.76 | 0.95 |
| **B* 07:02** | 0.74 | 0.76 | 1.00 | 0.75 | 0.76 | 1.00 |
| **B* 08:01** | 0.79 | 0.75 | 0.95 | 0.78 | 0.75 | 0.96 |
| **B* 13:02** | 0.86 | 0.75 | 0.98 | 0.86 | 0.75 | 0.98 |
| **B* 14:02** | 0.71 | 0.76 | 0.98 | 0.68 | 0.76 | 0.86 |
| **B* 15:01** | 0.83 | 0.75 | 0.85 | 0.79 | 0.75 | 0.98 |
| **B* 18:01** | 0.74 | 0.76 | 1.00 | 0.75 | 0.76 | 1.00 |
| **B* 27:05** | 0.86 | 0.75 | 0.62 | 0.86 | 0.75 | 0.63 |
| **B* 35:01** | 0.80 | 0.75 | 0.83 | 0.81 | 0.75 | 0.68 |
| **B* 35:03** | 0.71 | 0.76 | 1.00 | 0.71 | 0.76 | 1.00 |
| **B* 38:01** | 0.64 | 0.76 | 0.79 | 0.67 | 0.76 | 0.88 |
| **B* 39:01** | 0.83 | 0.75 | 0.96 | 0.83 | 0.75 | 0.96 |
| **B* 40:01** | 0.71 | 0.76 | 0.98 | 0.71 | 0.76 | 0.98 |
| **B* 40:02** | 0.75 | 0.75 | 1.00 | 0.75 | 0.75 | 1.00 |
| **B* 41:01** | 0.77 | 0.75 | 1.00 | 0.77 | 0.75 | 1.00 |
| **B* 44:02** | 0.75 | 0.76 | 1.00 | 0.76 | 0.75 | 1.00 |
| **B* 44:03** | 0.73 | 0.76 | 1.00 | 0.74 | 0.76 | 1.00 |
| **B* 49:01** | 0.59 | 0.76 | 0.40 | 0.59 | 0.76 | 0.41 |
| **B* 50:01** | 1.00 | 0.75 | 0.65 | 1.00 | 0.75 | 0.65 |
| **B* 51:01** | 0.82 | 0.75 | 0.79 | 0.79 | 0.75 | 0.94 |
| **B* 52:01** | 1.00 | 0.75 | 0.65 | 1.00 | 0.75 | 0.65 |
| **B* 53:01** | 0.83 | 0.75 | 1.00 | 0.83 | 0.75 | 1.00 |
| **B* 55:01** | 1.00 | 0.75 | 0.48 | 1.00 | 0.75 | 0.48 |
| **B* 57:01** | 0.71 | 0.76 | 0.95 | 0.74 | 0.76 | 1.00 |
| **B* 58:01** | 0.67 | 0.76 | 0.88 | 0.67 | 0.76 | 0.88 |
| **DQ* 02:01** | 0.78 | 0.75 | 0.93 | 0.77 | 0.75 | 0.99 |
| **DQ* 02:02** | 0.78 | 0.75 | 0.93 | 0.76 | 0.75 | 1.00 |
| **DQ* 03:01** | 0.80 | 0.72 | 0.39 | 0.80 | 0.74 | 0.41 |
| **DQ* 03:02** | 0.85 | 0.75 | 0.75 | 0.85 | 0.75 | 0.76 |
| **DQ* 03:03** | 0.79 | 0.75 | 1.00 | 0.82 | 0.75 | 0.90 |
| **DQ* 04:02** | 0.70 | 0.76 | 0.95 | 0.71 | 0.76 | 0.98 |
| **DQ* 05:01** | 0.72 | 0.77 | 0.81 | 0.70 | 0.76 | 0.47 |
| **DQ* 05:02** | 0.73 | 0.76 | 1.00 | 0.69 | 0.76 | 0.98 |
| **DQ* 05:03** | 0.80 | 0.75 | 1.00 | 0.80 | 0.75 | 1.00 |
| **DQ* 06:01** | 0.86 | 0.75 | 0.98 | 0.86 | 0.75 | 0.98 |
| **DQ* 06:02** | 0.67 | 0.78 | 0.18 | 0.65 | 0.77 | 0.11 |
| **DQ* 06:03** | 0.82 | 0.74 | 0.69 | 0.81 | 0.75 | 0.73 |
| **DQ* 06:04** | 0.83 | 0.75 | 1.00 | 0.83 | 0.75 | 1.00 |
| **DR* 01:01** | 0.74 | 0.76 | 1.00 | 0.72 | 0.76 | 0.89 |
| **DR* 01:02** | 0.57 | 0.76 | 0.78 | 0.50 | 0.76 | 0.44 |
| **DR* 03:01** | 0.77 | 0.75 | 1.00 | 0.76 | 0.75 | 1.00 |
| **DR* 03:02** | 0.83 | 0.75 | 1.00 | 0.83 | 0.75 | 1.00 |
| **DR* 04:01** | 0.67 | 0.76 | 0.63 | 0.67 | 0.76 | 0.65 |
| **DR* 04:02** | 0.86 | 0.75 | 0.84 | 0.86 | 0.75 | 0.84 |
| **DR* 07:01** | 0.75 | 0.76 | 1.00 | 0.74 | 0.76 | 0.97 |
| **DR* 08:01** | 0.67 | 0.76 | 0.88 | 0.69 | 0.76 | 0.94 |
| **DR* 09:01** | 0.83 | 0.75 | 0.96 | 0.83 | 0.75 | 0.96 |
| **DR* 10:01** | 0.75 | 0.75 | 1.00 | 0.75 | 0.75 | 1.00 |
| **DR* 11:01** | 0.80 | 0.75 | 0.83 | 0.81 | 0.75 | 0.64 |
| **DR* 11:04** | 0.75 | 0.75 | 1.00 | 0.75 | 0.75 | 1.00 |
| **DR* 12:01** | 1.00 | 0.75 | 0.35 | 1.00 | 0.75 | 0.35 |
| **DR* 13:01** | 0.67 | 0.78 | 0.28 | 0.65 | 0.77 | 0.17 |
| **DR* 13:02** | 0.64 | 0.76 | 0.85 | 0.64 | 0.76 | 0.85 |
| **DR* 13:03** | 0.90 | 0.74 | 0.43 | 0.90 | 0.75 | 0.45 |
| **DR* 14:54** | 0.78 | 0.75 | 1.00 | 0.78 | 0.75 | 1.00 |
| **DR* 15:01** | 0.75 | 0.76 | 1.00 | 0.75 | 0.75 | 1.00 |

| HBV | Phenotypic frequency | |  | Allelic frequency | |  |
| --- | --- | --- | --- | --- | --- | --- |
|  | Response |  |  | Response |  |  |
|  | **Positive** | **Negative** | **p-value** | **Positive** | **Negative** | **p-value** |
| A* 01:01 | 0.65 | 0.72 | 0.53 | 1.00 | 0.55 | 0.50 |
| A* 02:01 | 0.72 | 0.71 | 1.00 | 1.00 | 0.42 | 1.00 |
| A* 11:01 | 0.78 | 0.71 | 0.99 | 1.00 | 0.44 | 0.99 |
| A* 23:01 | 0.64 | 0.71 | 1.00 | 1.00 | 0.39 | 1.00 |
| A* 24:02 | 0.72 | 0.71 | 0.80 | 1.00 | 0.47 | 0.80 |
| A* 25:01 | 0.69 | 0.71 | 1.00 | 1.00 | 0.41 | 1.00 |
| A* 26:01 | 0.69 | 0.71 | 0.97 | 1.00 | 0.42 | 1.00 |
| A* 29:02 | 0.72 | 0.71 | 0.37 | 1.00 | 0.66 | 0.66 |
| A* 30:01 | 0.62 | 0.71 | 0.69 | 1.00 | 0.47 | 0.74 |
| A* 30:02 | 0.80 | 0.71 | 0.72 | 1.00 | 0.52 | 0.78 |
| A* 31:01 | 0.59 | 0.71 | 0.36 | 1.00 | 0.62 | 0.43 |
| A* 68:01 | 0.72 | 0.71 | 0.36 | 1.00 | 0.66 | 0.25 |
| A* 68:02 | 0.70 | 0.71 | 0.77 | 1.00 | 0.48 | 0.77 |
| A* 74:01 | 1.00 | 0.71 | 0.77 | 1.00 | 0.57 | 0.84 |
| B* 07:02 | 0.75 | 0.71 | 1.00 | 1.00 | 0.43 | 1.00 |
| B* 08:01 | 0.66 | 0.72 | 0.80 | 1.00 | 0.45 | 0.70 |
| B* 13:02 | 0.57 | 0.71 | 0.98 | 1.00 | 0.37 | 1.00 |
| B* 14:01 | 0.86 | 0.71 | 1.00 | 1.00 | 0.46 | 1.00 |
| B* 14:02 | 0.67 | 0.71 | 0.12 | 1.00 | 0.84 | 0.24 |
| B* 15:01 | 0.84 | 0.70 | 0.89 | 1.00 | 0.48 | 0.89 |
| B* 15:03 | 0.73 | 0.71 | 0.82 | 1.00 | 0.47 | 0.92 |
| B* 18:01 | 0.72 | 0.71 | 0.46 | 1.00 | 0.61 | 0.47 |
| B* 27:05 | 0.70 | 0.71 | 0.22 | 1.00 | 0.76 | 0.53 |
| B* 35:01 | 0.68 | 0.71 | 0.54 | 1.00 | 0.56 | 0.60 |
| B* 35:03 | 0.80 | 0.71 | 1.00 | 1.00 | 0.44 | 1.00 |
| B* 37:01 | 0.67 | 0.71 | 0.77 | 1.00 | 0.47 | 0.77 |
| B* 38:01 | 0.60 | 0.71 | 0.86 | 1.00 | 0.41 | 0.87 |
| B* 39:01 | 0.64 | 0.71 | 1.00 | 1.00 | 0.39 | 1.00 |
| B* 40:01 | 0.63 | 0.71 | 0.59 | 1.00 | 0.52 | 0.70 |
| B* 40:02 | 0.67 | 0.71 | 1.00 | 1.00 | 0.40 | 1.00 |
| B* 44:02 | 0.70 | 0.71 | 1.00 | 1.00 | 0.41 | 1.00 |
| B* 44:03 | 0.73 | 0.71 | 1.00 | 1.00 | 0.42 | 1.00 |
| B* 45:01 | 0.92 | 0.71 | 1.00 | 1.00 | 0.48 | 1.00 |
| B* 49:01 | 0.74 | 0.71 | 0.49 | 1.00 | 0.60 | 0.49 |
| B* 50:01 | 0.59 | 0.71 | 1.00 | 1.00 | 0.37 | 1.00 |
| B* 51:01 | 0.64 | 0.71 | 1.00 | 1.00 | 0.39 | 1.00 |
| B* 52:01 | 0.53 | 0.71 | 0.91 | 1.00 | 0.37 | 0.96 |
| B* 53:01 | 0.78 | 0.71 | 0.69 | 1.00 | 0.53 | 0.69 |
| B* 55:01 | 0.75 | 0.71 | 0.96 | 1.00 | 0.44 | 0.96 |
| B* 56:01 | 0.67 | 0.71 | 0.64 | 1.00 | 0.51 | 0.64 |
| B* 58:01 | 0.92 | 0.71 | 1.00 | 1.00 | 0.48 | 1.00 |
| DQ* 02:01 | 0.73 | 0.71 | 1.00 | 1.00 | 0.42 | 1.00 |
| DQ* 02:02 | 0.75 | 0.71 | 0.69 | 1.00 | 0.52 | 0.80 |
| DQ* 03:01 | 0.67 | 0.72 | 0.34 | 1.00 | 0.66 | 0.22 |
| DQ* 03:02 | 0.60 | 0.72 | 0.21 | 1.00 | 0.74 | 0.22 |
| DQ* 04:02 | 0.71 | 0.71 | 1.00 | 1.00 | 0.42 | 0.97 |
| DQ* 05:01 | 0.77 | 0.70 | 0.95 | 1.00 | 0.45 | 0.89 |
| DQ* 05:02 | 0.59 | 0.72 | 0.44 | 1.00 | 0.57 | 0.45 |
| DQ* 05:03 | 0.73 | 0.71 | 0.96 | 1.00 | 0.43 | 0.91 |
| DQ* 06:01 | 0.82 | 0.71 | 0.22 | 1.00 | 0.79 | 0.08 |
| DQ* 06:02 | 0.75 | 0.70 | 1.00 | 1.00 | 0.43 | 1.00 |
| DQ* 06:03 | 0.74 | 0.71 | 0.91 | 1.00 | 0.45 | 0.91 |
| DQ* 06:04 | 0.50 | 0.71 | 1.00 | 1.00 | 0.33 | 1.00 |
| DR* 01:01 | 0.78 | 0.70 | 0.81 | 1.00 | 0.49 | 0.80 |
| DR* 01:02 | 0.60 | 0.71 | 1.00 | 1.00 | 0.38 | 1.00 |
| DR* 03:01 | 0.73 | 0.71 | 1.00 | 1.00 | 0.42 | 0.89 |
| DR* 04:01 | 0.54 | 0.72 | 0.68 | 1.00 | 0.44 | 0.69 |
| DR* 04:02 | 0.55 | 0.72 | 0.66 | 1.00 | 0.46 | 0.38 |
| DR* 04:03 | 0.67 | 0.71 | 1.00 | 1.00 | 0.40 | 1.00 |
| DR* 04:04 | 0.76 | 0.71 | 0.98 | 1.00 | 0.44 | 1.00 |
| DR* 04:05 | 0.67 | 0.71 | 0.53 | 1.00 | 0.56 | 0.53 |
| DR* 07:01 | 0.71 | 0.71 | 0.09 | 1.00 | 0.89 | 0.20 |
| DR* 08:01 | 0.76 | 0.71 | 1.00 | 1.00 | 0.43 | 1.00 |
| DR* 09:01 | 0.75 | 0.71 | 1.00 | 1.00 | 0.43 | 1.00 |
| DR* 10:01 | 0.70 | 0.71 | 0.37 | 1.00 | 0.65 | 0.38 |
| DR* 11:01 | 0.67 | 0.71 | 0.66 | 1.00 | 0.50 | 0.58 |
| DR* 12:01 | 0.94 | 0.71 | 0.99 | 1.00 | 0.49 | 0.99 |
| DR* 13:01 | 0.72 | 0.71 | 0.91 | 1.00 | 0.44 | 0.91 |
| DR* 13:02 | 0.69 | 0.71 | 0.93 | 1.00 | 0.42 | 0.93 |
| DR* 13:03 | 0.88 | 0.71 | 0.90 | 1.00 | 0.49 | 0.90 |
| DR* 14:54 | 0.69 | 0.71 | 0.99 | 1.00 | 0.41 | 0.99 |
| DR* 15:01 | 0.76 | 0.70 | 1.00 | 1.00 | 0.43 | 1.00 |
| DR* 15:03 | 0.80 | 0.71 | 0.64 | 1.00 | 0.55 | 0.64 |
| DR* 16:01 | 0.52 | 0.72 | 0.98 | 1.00 | 0.35 | 0.94 |
| A* 01:01 | 0.65 | 0.72 | 0.53 | 1.00 | 0.55 | 0.50 |
| A* 02:01 | 0.72 | 0.71 | 1.00 | 1.00 | 0.42 | 1.00 |
| A* 11:01 | 0.78 | 0.71 | 0.99 | 1.00 | 0.44 | 0.99 |
| A* 23:01 | 0.64 | 0.71 | 1.00 | 1.00 | 0.39 | 1.00 |
| A* 24:02 | 0.72 | 0.71 | 0.80 | 1.00 | 0.47 | 0.80 |
| A* 25:01 | 0.69 | 0.71 | 1.00 | 1.00 | 0.41 | 1.00 |
| A* 26:01 | 0.69 | 0.71 | 0.97 | 1.00 | 0.42 | 1.00 |
